# Epigenetic age and socioeconomic status contribute to racial disparities in cognitive and functional aging between Black and White older Americans

**DOI:** 10.1101/2023.09.29.23296351

**Authors:** Isabel Yannatos, Shana D. Stites, Courtney Boen, Sharon X. Xie, Rebecca T. Brown, Corey T. McMillan

**Author notes:** Address correspondence to: Corey T. McMillan, PhD.

## Abstract

Epigenetic age, a biological aging marker measured by DNA methylation, is a potential mechanism by which social factors drive disparities in age-related health. Epigenetic age gap is the residual between epigenetic age measures and chronological age. Previous studies showed associations between epigenetic age gap and age-related outcomes including cognitive capacity and performance on some functional measures, but whether epigenetic age gap contributes to disparities in these outcomes is unknown. We use data from the Health and Retirement Study to examine the role of epigenetic age gap in racial disparities in cognitive and functional outcomes and consider the role of socioeconomic status (SES). Epigenetic age measures are GrimAge or Dunedin Pace of Aging methylation (DPoAm). Cognitive outcomes are cross-sectional score and two-year change in Telephone Interview for Cognitive Status (TICS). Functional outcomes are prevalence and incidence of limitations performing Instrumental Activities of Daily Living (IADLs). We find, relative to White participants, Black participants have lower scores and greater decline in TICS, higher prevalence and incidence rates of IADL limitations, and higher epigenetic age gap. Age- and gender-adjusted analyses reveal that higher GrimAge and DPoAm gap are both associated with worse cognitive and functional outcomes and mediate 6-11% of racial disparities in cognitive outcomes and 19-39% of disparities in functional outcomes. Adjusting for SES attenuates most DPoAm associations and most mediation effects. These results support that epigenetic age gap contributes to racial disparities in cognition and functioning and may be an important mechanism linking social factors to disparities in health outcomes.

## Introduction

Racial disparities in cognitive performance and instrumental activities of daily living (IADL) impairments are well documented. Performance of IADLs, activities including managing money and taking medications, is important for independent living and tends to decline with age. Black older Americans are consistently found to have lower scores on cognitive tests (1,2) and higher rates of IADL impairments (3–10). Studies of disparities in longitudinal change in cognitive scores have found mixed results (1,2), while studies of IADL limitations over time show earlier onset and steeper rates of decline for Black compared to White adults (5,8–10). It is important to understand how race – a social construct – leads to biological differences in cognitive and functional aging. The weathering hypothesis is a prominent explanation for racialized differences in the pace of age-related health decline (11).

Weathering refers to accelerated biological aging for Black Americans and other marginalized populations relative to White individuals because of the cumulative impacts of racism, discrimination, and structural disadvantage (12). In recent years, scholars have used markers of biological aging to directly test the weathering hypothesis. Measures of biological aging are meant to capture heterogeneity in the rate of physiological change with chronological aging and predict age-related health outcomes. A growing body of research documents racial disparities in biological aging using several methods, including epigenetic age based on DNA methylation (DNAm) (13,14). As a result, epigenetic age has gained attention as a potential mechanism underlying racial disparities across a range of health outcomes (15–17). However, it is not known whether epigenetic age measures are associated with age-related functional outcomes, such as ability to perform IADLs, nor whether they contribute to racial disparities in cognitive or functional aging.

We and others demonstrated large racial disparities in GrimAge and Dunedin Pace of Aging methylation (DPoAm) gaps (13,14), two prominent measures of epigenetic age that capture phenotypic age based on biomarkers. Older epigenetic age than chronological age, or a positive epigenetic age gap, indicates accelerated biological aging. Black individuals have higher average epigenetic age gap than White individuals. Both GrimAge and DPoAm use DNAm, a reversible chemical modification of DNA, to capture phenotypic changes associated with aging by selecting a set of DNAm CpG sites that are highly associated with blood biomarkers of aging and function (18,19). GrimAge and DPoAm are associated with and contribute to disparities in many age-related outcomes (13).

Previous studies showed associations between both GrimAge and DPoAm and cognitive test outcomes, but it is unknown whether they contribute to racial disparities in these outcomes (18–24). GrimAge and DPoAm have also previously been associated with some physical function outcomes, such as gait speed and balance, and basic activities of daily living (BADLs): ability to dress, eat, bathe, toilet, and transfer independently (13,18,19,22,24,25). Racial disparities in GrimAge and DPoAm gaps were shown to contribute significantly to racial disparities in the number of physical and BADL limitations and change in the number of BADL limitations (13). There are no studies to our knowledge examining associations between epigenetic age gap and IADL outcomes, which have a higher cognitive demand than physical and BADL function.

Further, the role of socioeconomic status (SES) in these links remains to be better understood. SES also has a large influence on cognitive and functional aging and contributes to disparities in these outcomes. Education and financial resources shape one’s opportunities, environments, experiences, and health; inequitable distribution of these resources is a fundamental pathway by which racism operates to produce health disparities (26). There are socioeconomic gradients in aging, including cognitive scores (27–29), IADL limitations (5,6,9), and epigenetic age gap (30,31), where these outcomes are strongly associated with SES. Still, many previous analyses either do not consider the role of SES or include only one indicator such as income when examining associations between epigenetic age and health (13,18–22,25). While education, income, and wealth are related, they each reflect different aspects of SES and have separate effects on aging (27). There is evidence that GrimAge and DPoAm mediate the association between SES (as an index of education and wealth) and cognitive test scores (23), leading to the question of whether epigenetic age gap has an effect independent of SES on cognitive and functional aging. We address this by examining models with and without education and an index of wealth and income.

In this study we investigate the associations between epigenetic age gap and cognitive and functional outcomes, the contribution of Black-White racial disparities in epigenetic age gap to disparities in these outcomes, and the role of SES in these relationships. We hypothesize that higher GrimAge and DPoAm gaps are associated with worse outcomes and mediate significant portions of the racial disparities in these outcomes, with SES playing a significant role in these associations. We test these hypotheses using a large, well-characterized, nationally representative study sample and two robust measures of epigenetic age. This study expands understanding of the role of weathering - as measured by multiple epigenetic age measures - in generating racial disparities in age-related cognition and function.

## Methods

### Data

The Health and Retirement Study (HRS) is sponsored by the National Institute on Aging (grant number NIA U01 AG009740) and conducted by the University of Michigan. It is a longitudinal panel survey study, administering surveys biannually to a nationally representative population of Americans aged 50 or older (32). A subset of HRS participants in the 2016 wave provided a venous blood sample, a representative subsample of which (N=4104) was selected for DNA methylation (DNAm) measurement (33). We link epigenetic DNAm age data with survey responses from 2014-2018, cognitive test data from 2016-2018, and a cross-wave tracker file that includes sample weights.

### Population

We include 3,282 participants in the HRS DNAm subsample who self-identify as non-Hispanic White (N=2,636) or non-Hispanic Black (N=646). Inclusion criteria are having a DNAm measurement that passed quality control (N=4018) and identifying as non-Hispanic and White or Black (N=3326). Exclusion criteria are missing demographic or socioeconomic status (SES) data (education, wealth or income, N=44).

### Measures

#### Outcomes

We include two outcome measures and for each examine one cross-sectional outcome in 2016, the year DNAm was measured, and one outcome reflecting change between the 2016 and 2018 HRS waves. All participants have complete outcome data in 2016; those missing 2018 outcome data are excluded from respective longitudinal analyses. First, we use the modified Telephone Interview for Cognitive Status (TICS) (34,35) that includes items for memory (immediate and delayed recall), working memory (serial 7 subtraction test), and attention and processing speed (backward count test) for a total of 27 points. The modified version excludes orientation and naming items which were only collected from HRS participants older than 65 years (36). We use TICS score in 2016 as a cross-sectional outcome for all participants and the difference in TICS score between 2016 and 2018 as a longitundinal outcome for 2,849 participants, excluding 433 participants missing 2018 TICS data.

Second, we use Instrumental Activities of Daily Living (IADL) limitations as a functional outcome measure. The HRS survey asks whether respondents have any difficulty with five tasks (using the phone, managing money, taking medications, shopping for groceries, and preparing hot meals). Any difficulty in any IADL task is considered an IADL limitation. We use prevalence (presence or absence) of any IADL limitation in 2016 as a cross-sectional outcome. We use incidence (among those without a prevalent limitation, whether they develop a new limitation) in 2018 as a longitudinal outcome in 2,538 participants, excluding 439 participants who have a prevalent IADL limitation and an additional 305 missing 2018 IADL data.

#### Epigenetic age gap

Collection and measurement of DNAm samples during the 2016 HRS wave have been previously described (33). Whole blood samples were collected in EDTA tubes and sent to the CLIA-certified Advanced Research and Diagnostic Laboratory at the University of Minnesota for processing (33). DNAm was measured using the Infinium Methylation EPIC BeadChip.

Samples were run in duplicate, randomized across plates, and quality controlled. Epigenetic age values, including those for GrimAge and Dunedin Pace of Aging methylation (DPoAm), were constructed based on published algorithms that combine and weight methylation levels at specific sites (18,19). For both GrimAge and DPoAm, we regress the epigenetic age value on chronological age so the residual represents epigenetic age gap. We then scale for ease of comparison between the two measures by dividing by the root mean square. A positive residual indicates accelerated epigenetic age gap, or higher epigenetic age than expected based on chronological age.

#### Covariates

In all statistical models we adjust for age, self-reported race (White or Black), and self-reported gender. In the HRS survey there is no distinction between assigned sex or gender identity. Gender is treated as a binary and “sex-mismatched” blood samples were removed from DNAm measurement. In statistical models referred to as “SES models”, we additionally adjust for education and wealth/income. Education is based on years of formal schooling completed and is categorized as less than high school (<12 years), high school (12 years), some college (13-15 years), or college or more (≥16 years). Wealth/income is divided into weighted quartiles based on an index of 2014 household wealth and income. Household wealth is the sum of assets, including second homes, minus debts. Household income is the total of respondent and spouse annual incomes, divided by the square root of number of individuals in the household. We use log-transformed values for both because of skew in the distributions, calculate Z scores for each, take the average of the Z scores, and classify into weighted quartiles. The first quartile represents the lowest level of wealth/income and the fourth represents the highest.

We conduct supplemental models including health behavior and health status variables that are potential confounders of epigenetic age and cognitive functioning as additional covariates. Health behavior variables include tobacco smoking, alcohol consumption, and frequency of physical activity. Health status variables include self-rated health, number of depression symptoms, number of chronic conditions, and history of stroke. These variables are commonly included in the aging and disparities literature and were found to be significantly associated with both epigenetic age gap and at least one outcome measure in univariate analyses. See Supplement for details.

#### Sample weights

To adjust for potential sources of bias in the probability of participation in the DNAm subsample (33), sample weights are provided by HRS. We impute missing sample weights (N=143 missing, 4.4%) using the mean value and include weighting in all analyses. For mediation analyses it is necessary to scale the weights such that the sum of sample weights is equal to the sample size.

### Analytic approach

All analyses are conducted in R statistical software (version 4.2.1) and code is available at github.com/pennbindlab (37). Models are informed by a directed acyclic graph showing proposed relationships between all measures (Fig 1). All models include race, age, gender, and sample weights. SES is both a confounder of the epigenetic age gap-outcome associations and a mediator on the pathway between race and outcomes. Models with the TICS change outcome are also adjusted for baseline TICS score in 2016, since magnitude of the baseline score may influence the amount of possible change. We use linear models for the continuous TICS outcomes and logistic models for the binary IADL outcomes. All statistical tests were two-sided. Statistical significance was set at p<0.05. We use two modeling approaches to test our hypotheses: regression and mediation.

**Fig 1.**
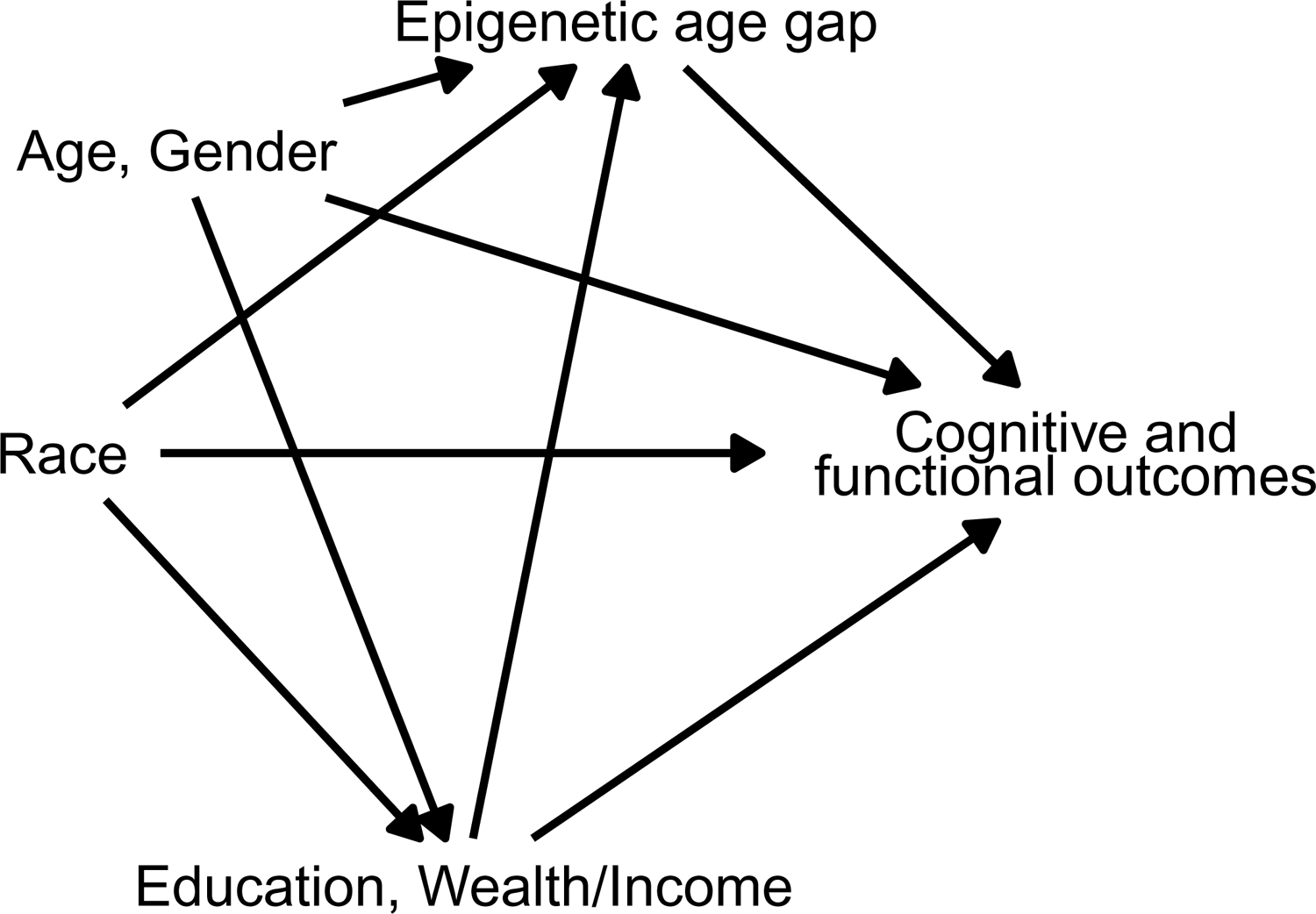
Directed acyclic graph showing relationships between variables. Through various forms of racism, race is associated with SES (education, income, and wealth), epigenetic age gap, and with cognitive and functional outcomes as evidenced by racial disparities in these measures. Age, gender, and SES are associated with both epigenetic age gap and these outcomes. We hypothesize an association between epigenetic age gap and both cognitive and functional outcomes.

### Regression models

We use a stepwise approach to investigate the associations between epigenetic age gap, race, and SES and each cognitive and functional outcome. First, we examine models that adjust for only race, age, and gender, which we refer to as “basic models”. Second, we add education and the wealth/income index to what we refer to as “SES models”. Third, we add each epigenetic age gap measure separately to both basic and SES models. Variance inflation factors for all models are ≤1.2, indicating low collinearity of the explanatory variables. Logistic models were implemented using the survey package to correctly account for weights (38). P-values are Bonferroni corrected to account for four comparisons. AIC (Aikake’s Information Criterion) is shown as a goodness-of-fit indicator. We conduct supplemental models to assess the confounding effects of health behavior and health status variables (see Supplement).

### Mediation models

Next, we implement mediation analyses to quantify the extent to which the racial disparity in each epigenetic age gap measure contributes to racial disparities in each outcome. In these analyses, race is the exposure and epigenetic age gap is the mediator. We use a *g* formula approach as implemented in the CMAVerse package because it allows inclusion of SES as a mediator-outcome confounder that is affected by the exposure (39). Including SES as a regular covariate would violate the assumption in mediation that there are no mediator-outcome confounders that are affected by the exposure, since SES is profoundly affected by race, yet including it as an additional mediator would not allow us to separate the effects of SES and epigenetic age gap from their joint effect. Similar to the regression approach, we first examine basic models that include only age and gender as confounders, then add education and wealth/income to SES models as post-exposure confounders. Models with the TICS change outcome adjust for baseline TICS score by using the residual of TICS change from TICS score in 2016 as the outcome. All models include scaled sample weights.

## Results

Descriptive characteristics of the study population by race are shown in Table 1. Overall, the sample is 71 years old on average, 42% male, and 80.3% White. Pairwise comparisons demonstrate that White participants are significantly older and include a higher proportion of men than Black participants, and have significantly higher TICS scores, lower likelihood of a prevalent IADL limitation, lower epigenetic age gaps, higher likelihood of having college education, and higher likelihood of having above-median wealth/income. There is no significant difference in the change in TICS score or the incidence of IADL limitations between White and Black participants.

**Table 1.** Sample characteristics by race.

| **Characteristic** | **Overall, N = 3,282<sup>1</sup>** | **White, N = 2,636<sup>1</sup>** | **Black, N = 646<sup>1</sup>** | **p-value<sup>2</sup>** | **Effect Size<sup>3</sup>** |
| --- | --- | --- | --- | --- | --- |
| **Age** | 70.77 (9.65) | 71.68 (9.68) | 67.06 (8.57) | <0.001 | 0.506 |
| **Gender** |  |  |  | <0.001 | 0.078 |
| Male | 1,371 (42%) | 1,152 (44%) | 219 (34%) |  |  |
| Female | 1,911 (58%) | 1,484 (56%) | 427 (66%) |  |  |
| **TICS score** | 15.27 (4.29) | 15.69 (4.15) | 13.55 (4.44) | <0.001 | 0.499 |
| **Change in TICS score** | 0.14 (3.72) | 0.16 (3.68) | 0.09 (3.87) | 0.7 | 0.018 |
| Unknown | 433 | 345 | 88 |  |  |
| **IADL limitation prevalence** | 439 (13%) | 333 (13%) | 106 (16%) | 0.012 | 0.043 |
| **IADL limitation incidence** | 203 (8.0%) | 156 (7.5%) | 47 (10%) | 0.076 | 0.033 |
| Unknown | 744 | 568 | 176 |  |  |
| **GrimAge gap** | 0.06 (1.02) | -0.01 (1.00) | 0.33 (1.06) | <0.001 | 0.332 |
| **DPoAm gap** | 0.06 (1.01) | -0.01 (0.98) | 0.35 (1.07) | <0.001 | 0.355 |
| **Education** |  |  |  | <0.001 | 0.159 |
| College + | 917 (28%) | 796 (30%) | 121 (19%) |  |  |
| Some College | 862 (26%) | 693 (26%) | 169 (26%) |  |  |
| High School | 1,065 (32%) | 859 (33%) | 206 (32%) |  |  |
| < High School | 438 (13%) | 288 (11%) | 150 (23%) |  |  |
| **Wealth/Income Quartile** |  |  |  | <0.001 | 0.380 |
| 4 | 748 (23%) | 722 (27%) | 26 (4.0%) |  |  |
| 3 | 827 (25%) | 749 (28%) | 78 (12%) |  |  |
| 2 | 877 (27%) | 695 (26%) | 182 (28%) |  |  |
| 1 | 830 (25%) | 470 (18%) | 360 (56%) |  |  |

<sup>1</sup>Mean (SD); n (%)
<sup>2</sup>Wilcoxon rank sum test; Pearson's Chi-squared test
<sup>3</sup>Cohen's D; Cramer's V

### Regression models: association of epigenetic age gap with cognitive and functional outcomes

The magnitude of the regression coefficients for GrimAge and DPoAm gaps and each outcome are shown in Figure 2. In basic models, both GrimAge and DPoAm gaps are significantly associated with all outcomes. Higher epigenetic age gap is associated with lower TICS score, greater decline in TICS change, and higher odds of both IADL limitation prevalence and incidence. The magnitude of associations with all outcomes is larger for GrimAge than DPoAm and larger for TICS score than TICS change. Full results of TICS score models are shown in eTable 1; coefficient values are GrimAge β = −0.77, DPoAm β = −0.47. TICS change models are shown in eTable 2; coefficient values are GrimAge β = −0.42, DPoAm β = −0.26.

**Fig 2.**
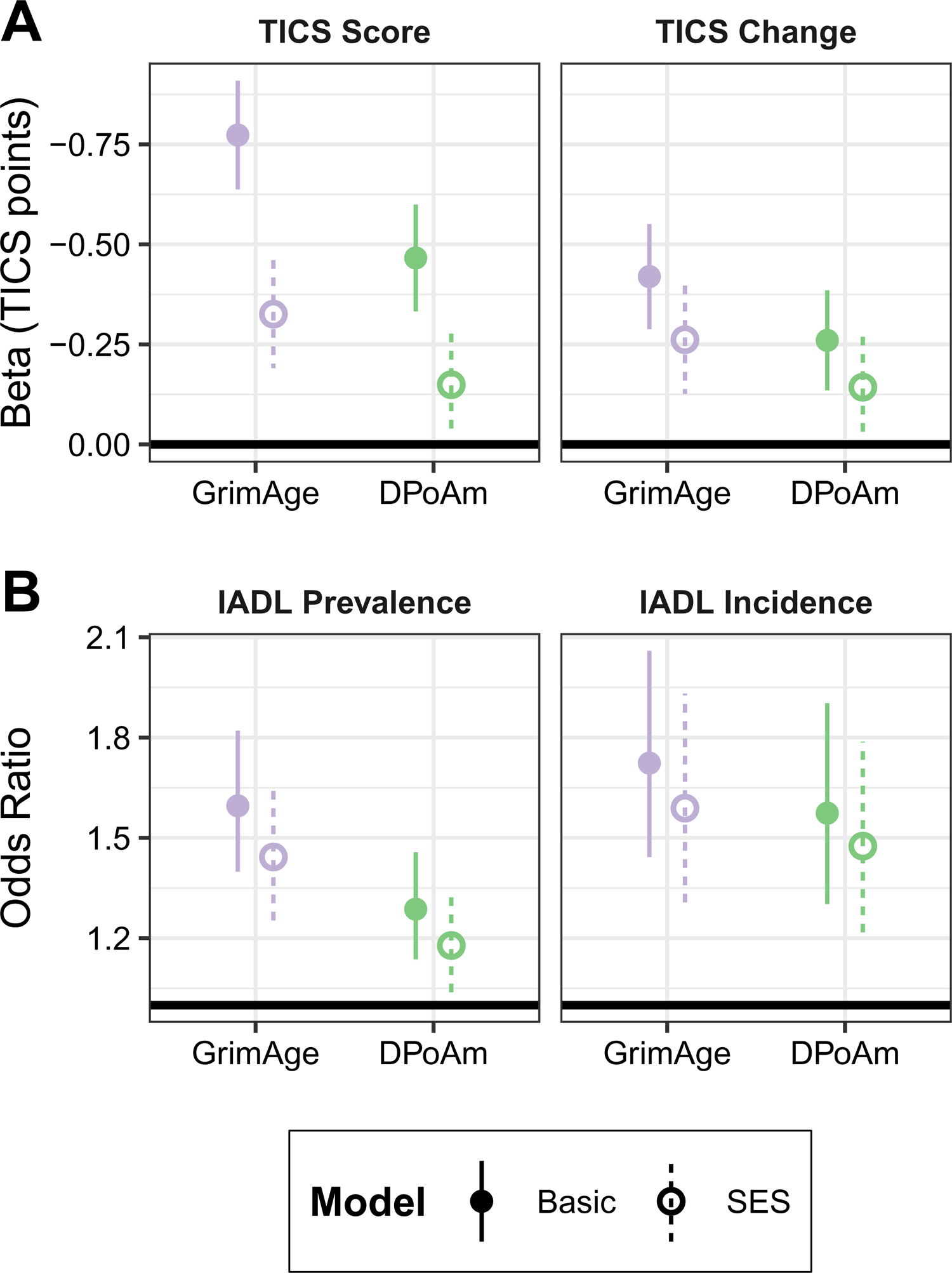
Associations between epigenetic age gap and outcomes. Points represent the magnitude of the association and lines represent 95% confidence intervals. Basic models, adjusted for race, age, and gender, are shown with solid points and lines; SES models, which include education and wealth/income, are shown with empty points and dashed lines. The coefficients of GrimAge and DPoAm gaps with outcomes A) TICS score and TICS change and B) IADL limitation prevalence and IADL limitation incidence are plotted.

The magnitude of associations with IADL limitation incidence are larger than with IADL limitation prevalence. IADL limitation prevalence models are shown in eTable 3; coefficient values are GrimAge odds ratio (OR) = 1.6, DPoAm OR = 1.29. IADL limitation incidence models are shown in eTable 4; coefficient values are GrimAge OR = 1.72, DPoAm OR = 1.57.

Adding education and wealth/income variables in SES models reduces the magnitudes of all epigenetic age gap and outcome associations (Fig 2). GrimAge gap remains associated with all outcomes while DPoAm gap is associated only with IADL limitation incidence. Coefficient values for TICS score are GrimAge β = −0.33, DPoAm β = −0.15. Coefficient values for TICS change are GrimAge β = −0.26, DPoAm β = −0.14. Coefficient values for IADL limitation prevalence are GrimAge OR = 1.44, DPoAm OR = 1.18. Coefficient values for IADL limitation incidence are GrimAge OR = 1.59, DPoAm OR = 1.48.

Supplemental models include health behavior and health status variables as additional covariates. Descriptive summaries of these variables are shown in eTable 5. Results of regression models including these variables are shown in eTables 6 and 7. After adding both health behaviors and health status to SES models, GrimAge gap remains associated with both TICS score (β = −0.25) and TICS change (β = −0.22). DPoAm gap remains not associated with either TICS outcome. Adding health behavior variables to SES models has little impact on IADL associations; GrimAge gap remains associated with both IADL limitation prevalence (OR = 1.36) and incidence (OR = 1.56), while DPoAm gap remains associated only with IADL limitation incidence (OR = 1.41). Adding health status variables attenuates the association between GrimAge gap with IADL limitation prevalence (OR = 1.20), but both GrimAge and DPoAm gaps remain associated with IADL limitation incidence (GrimAge OR = 1.48, DPoAm OR = 1.36).

### Regression models: magnitude of racial disparities in outcomes

We examine the race coefficient in regression models to determine the racial disparities in each outcome and the contributions of SES and epigenetic age gap to these disparities (Figure 3). In basic models adjusting for only age and gender, without epigenetic age gap, there are significant racial disparities in all outcomes. Black participants have 3.06 points lower TICS scores, 1.79 points more decline in TICS score, 2.01 times higher odds of prevalent IADL limitation, and 2.02 times higher odds of incident IADL limitation than White participants. Adding SES to the models decreases the magnitude of disparities in all outcomes and attenuates the disparities in IADL limitation prevalence and incidence to non-significance. Adding GrimAge or DPoAm gap to either the basic or SES model decreases the magnitude of the racial disparities but has a much smaller effect than adding SES to the basic model.

**Fig 3.**
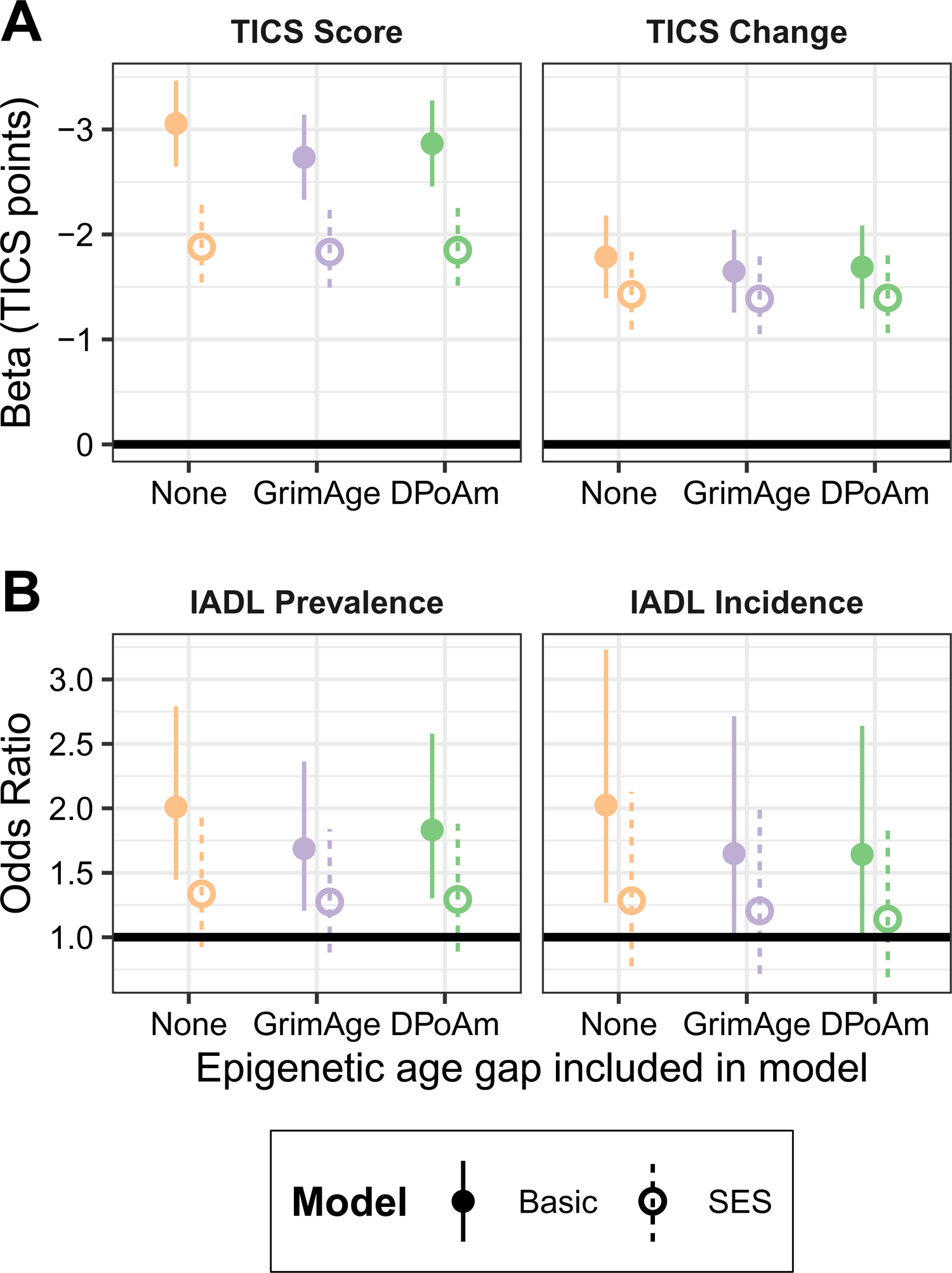
Racial disparity in outcomes. Points represent the magnitude of the race coefficient (White being the reference group) and lines represent 95% confidence intervals. Basic models, adjusted for age and gender, are shown with solid points and lines; SES models, which include education and wealth/income, are shown with empty points and dashed lines. The race coefficients with GrimAge gap, DPoAm gap, or neither are shown for A) TICS score and TICS change, B) IADL limitation prevalence and incidence.

### Mediation models: contribution of epigenetic age gap to disparities in outcomes

Results of mediation models with race as the exposure, epigenetic age gap as the mediator, and each of the four outcomes are shown in Table 2. In basic models adjusting for age and gender, both GrimAge and DPoAm gaps mediate significant portions of the racial disparities in all outcomes. GrimAge gap mediates 10.5% of the disparity in TICS score, 10.52% of the disparity in TICS change, 33.71% of the disparity in IADL limitation prevalence, and 39.08% of the disparity in IADL limitation incidence. DPoAm gap mediates 6.19% of the disparity in TICS score, 6.75% of the disparity in TICS change, 18.83% of the disparity in IADL limitation prevalence, and 38.51% of the disparity in IADL limitation incidence.

**Table 2.** Mediating effects of epigenetic age gap on racial disparities in outcomes.

| Mediator: |  | GrimAge |  | DPoAm |  |
| --- | --- | --- | --- | --- | --- |
| Model: |  | Basic | SES | Basic | SES |
| Outcome | Effect |  |  |  |  |
| TICS Score | Indirect | -0.32***<br>(-0.43, -0.21) | -0.06***<br>(-0.13, -0.03) | -0.19***<br>(-0.28, -0.11) | -0.44<br>(-1.03, 0.73) |
|  | Total | -3.06***<br>(-3.49, -2.58) | -2.17***<br>(-2.63, -1.68) | -3.06***<br>(-3.47, -2.70) | -2.60***<br>(-3.13, -1.31) |
|  | Mediated | **0.11\*\*\***<br>**(0.07, 0.14)** | **0.03\*\*\***<br>**(0.01, 0.06)** | **0.06\*\*\***<br>**(0.04, 0.09)** | 0.17<br>(-0.54, 0.36) |
| TICS Change | Indirect | -0.16***<br>(-0.26, -0.08) | 0.20<br>(-1.05, 1.10) | -0.10***<br>(-0.16, -0.02) | -0.29<br>(-0.78, 0.67) |
|  | Total | -1.48***<br>(-1.83, -1.10) | -0.86<br>(-2.12, 0.35) | -1.48***<br>(-1.89, -1.11) | -1.37*<br>(-1.94, -0.32) |
|  | Mediated | **0.11\*\*\***<br>**(0.05, 0.18)** | -0.23<br>(-2.52, 3.21) | **0.07\*\*\***<br>**(0.01, 0.11)** | 0.21<br>(-1.31, 0.49) |
| IADL Prevalence | Indirect | 1.20***<br>(1.13, 1.27) | 1.07***<br>(1.03, 1.14) | 1.10***<br>(1.05, 1.16) | 1.04*<br>(1.01, 1.08) |
|  | Total | 1.96***<br>(1.35, 2.60) | 1.29<br>(0.90, 1.84) | 1.97***<br>(1.45, 2.63) | 1.31<br>(0.87, 1.87) |
|  | Mediated | **0.34\*\*\***<br>**(0.23, 0.58)** | 0.31<br>(-0.90, 2.09) | **0.19\*\*\***<br>**(0.09, 0.32)** | 0.16<br>(-0.83, 1.18) |
| IADL Incidence | Indirect | 1.24***<br>(1.15, 1.37) | 1.12***<br>(1.05, 1.23) | 1.24*<br>(1.11, 1.38) | 1.15***<br>(1.06, 1.27) |
|  | Total | 2.01***<br>(1.21, 3.03) | 1.34<br>(0.80, 2.09) | 1.99*<br>(1.17, 2.99) | 1.34<br>(0.73, 2.25) |
|  | Mediated | **0.39\*\*\***<br>**(0.23, 1.11)** | 0.43<br>(-1.51, 10.44) | **0.39\***<br>**(0.18, 0.85)** | 0.50<br>(-2.78, 2.65) |

Note. The indirect and total effects and the proportion mediated are shown with 95% confidence intervals. Units of the effects are TICS points for TICS outcomes and odds ratio for IADL outcomes. Proportion mediated is a relative proportion for all outcomes.
\*p<0.05; \*\*p<0.01; \*\*\*p<0.001

Adding education and wealth/income as post-exposure confounders attenuates mediating effects for TICS outcomes. Only the mediating effect of GrimAge gap for TICS score remains significant (2.76%). For IADL outcomes, adding SES attenuates the magnitude of the racial disparities, so the total effect of race is no longer significant.

## Discussion

We investigate the relationships of epigenetic age gap with cognitive performance and instrumental activities of daily living (IADL) functioning using both cross-sectional and longitudinal measures. We find that higher epigenetic age gap, measured by either GrimAge or Dunedin Pace of Aging methylation (DPoAm), is associated with lower scores and steeper decline on cognitive testing, and greater risk of IADL limitations. We also find that epigenetic age gap partially mediates the association between race and these outcomes, indicating that the Black-White racial disparity in epigenetic age gap contributes to disparities in cognitive and functional outcomes. Socioeconomic status (SES) also contributes to racial disparities in these outcomes and plays an important role in the pathway from race to epigenetic age gap and age-related outcomes. These results extend previous findings on physical aging to cognitive and functional aging, showing that GrimAge and DPoAm gaps are associated with and contribute to disparities in both.

We find some notable differences between the GrimAge and DPoAm epigenetic age gap measures. GrimAge gap has larger associations than DPoAm gap with all outcomes examined. GrimAge gap also remains associated with the outcomes after adjusting for SES variables and health behavior variables, such as tobacco and alcohol consumption and physical activity. After adding health status variables, such as self-rated health and chronic conditions, GrimAge gap remains associated with all outcomes except for IADL limitation prevalence. This demonstrates the strong association of GrimAge gap with cognitive and functional outcomes accounting for many confounding factors. In contrast, DPoAm gap remains associated with only IADL limitation incidence after adjusting for SES and health variables. In mediation analyses, GrimAge gap mediates a larger portion of disparities in all outcomes than DPoAm gap. This is likely due to the stronger association of GrimAge gap with the outcomes rather than a stronger association with race, since we previously found a stronger association of race with DPoAm gap than with GrimAge gap (14).

These findings are consistent with previous reports that GrimAge is more strongly associated than DPoAm with cognitive and physical function outcomes (13,23). GrimAge may be a more robust predictor of age-related health in general. Several differences in the construction of these measures could account for this. First, GrimAge was constructed using data from the Framingham Heart Study Offspring cohort which has an average age of approximately 70 years at the time of DNA methylation (DNAm) data collection, whereas DPoAm was constructed using data from a birth cohort study at ages 26-38 (18,19). The older and wider age range of participants for GrimAge may contribute to it being more sensitive to later-life aging processes. Second, DPoAm is constructed as a combination of DNAm levels at only 46 sites, while GrimAge combines 1030 sites. This may make DPoAm more subject to measurement error and variability, biasing its associations downward. A more recent version, called DunedinPACE (Pace of Aging Calculated from the Epigenome) addresses this limitation by using only DNAm sites with high test-retest reliability and includes 173 DNAm sites (40). This version demonstrates improved reliability and stronger associations with age-related outcomes compared to the original DPoAm and to GrimAge (24,40). We were not able to examine DunedinPACE due to data availability; future studies should incorporate and make available next-generation epigenetic aging measures such as DunedinPACE. A third difference between these measures is that GrimAge estimates static biological age while DPoAm is meant to estimate the rate of biological aging per chronological year. Results showing more similar associations between GrimAge and DPoAm for the longitudinal outcomes compared to cross-sectional outcomes are consistent with this idea.

We also find differences between cross-sectional and longitudinal outcomes that are not consistent across Telephone Interview for Cognitive Status (TICS) and IADL outcomes. Epigenetic age gap is more strongly associated with cross-sectional TICS score than longitudinal TICS change, consistent with previous findings (22,23). This could reflect that cognitive testing performance is more related to biological aging than is rate of decline, or it could reflect limitations in the measurement of cognitive decline. TICS has limited sensitivity and precision and we have only two time points. In contrast, epigenetic age gap is more weakly associated with cross-sectional IADL limitation prevalence than with longitudinal limitation incidence. Epigenetic age gap also mediates a smaller portion of the racial disparity in IADL limitation prevalence than incidence. Having a prevalent IADL limitation may be related to long-term disability or illness and less related to biological aging compared to a limitation that develops in later life.

Epigenetic age gap mediates a substantial portion of the racial disparity in IADL limitation prevalence (34% and 19% for GrimAge and DPoAm, respectively). Comparing our mediation results to those of a similar analysis by Graf *et al* shows that the contribution of epigenetic age gap to the disparity in prevalence of IADL limitations is similar to its contribution to racial disparities in prevalence of physical and basic activities of daily living (BADL) limitations (13). GrimAge and DPoAm mediated 43% and 34%, respectively, of the racial disparity in physical functional impairment and 16% and 13% of the disparity in BADL impairment (13). We cannot directly compare the mediation results with longitudinal outcomes since Graf *et al* examine change in the number of limitations while we examine incidence of new limitations. We find that epigenetic age gap mediates a large portion of the disparity in IADL limitation incidence (39%).

Epigenetic age gap mediates a significant but smaller portion of disparities in TICS score (11% and 6% for GrimAge and DPoAm, respectively) and change in TICS score (11% and 7%). This may reflect that both GrimAge and DPoAm were developed to predict blood biomarkers, which may be more closely linked to physical function than to cognitive aging. It may also reflect that factors besides aging make a larger contribution to disparities in cognitive test performance. The participants included in this study were born in 1966 and earlier, with the average year of birth in 1945. Many Black Health and Retirement Study (HRS) participants have early life experiences of lower quality of education, segregation, and discrimination which can contribute to less developed test-taking skills and cognitive reserve than their White counterparts, putting them at greater risk of scoring lower on neuropsychological tests regardless of age (2,27). There is also evidence of measurement variance in TICS and similar neuropsychological tests, where the measures may not function the same across racial groups (41–43).

Our study rigorously and innovatively examines the role of SES in these relationships by including education, income, and wealth data in our analyses. The results support a strong contribution of SES to racial disparities in cognition and functioning. Education (based on years of schooling) and household wealth/income quartile are significantly associated with all outcomes. Adding these variables to models with GrimAge and DPoAm gap reduces the magnitude of all associations between epigenetic age gap and outcomes. We and others have previously found strong associations between both GrimAge and DPoAm with SES measures (14,30). It is important to include these measures as confounders when examining the associations between epigenetic aging and health outcomes. Health behavior and health status variables have weaker attenuating effects; adding these variables decreases associations of epigenetic age gap with outcomes to a much smaller extent than adding SES variables. The exception is the attenuation of epigenetic age gap associations with IADL limitation prevalence upon adding health variables; this may be due to reverse causation. A prevalent IADL limitation due to long-term disability or illness may lead to decreased physical activity, poorer self-rated health, depressive symptoms, and chronic conditions.

Inequitable levels of education, wealth, and income between Black and White Americans greatly contribute to disparities in age-related health outcomes (11). We find that accounting for SES substantially reduces the magnitude of racial disparities in TICS outcomes and brings disparities in IADL outcomes to non-significance. Other studies have also found large contributions of SES to disparities in cognitive and functional outcomes (9,28,44–47). In mediation analyses, after adding SES variables the total effect of race on IADL outcomes is non-significant while the indirect effects of epigenetic age gap remain significant. For TICS outcomes, the indirect effects of epigenetic age gap become insignificant except for GrimAge with TICS score. GrimAge gap has a small contribution to the disparity in TICS score (3%) independent of SES. These results are largely consistent with the role of epigenetic age gap as mainly a downstream mediator of SES and a potential mechanism by which SES is biologically embedded to produce racial disparities in aging-related health. We demonstrated in prior work that inequitable levels of SES also contribute substantially to the racial disparity in epigenetic age gap (14). Disparate levels of education explained 11% and 7% of the disparity in GrimAge and DPoAm, respectively, and inequitable distribution of wealth/income explained 21% and 14%. There is evidence that GrimAge and DPoAm mediate significant portions of the associations between SES and cognitive test performance and decline, but this hypothesis has not been directly tested for IADL outcomes (23).

Taken together, these results support the weathering hypothesis and the role of epigenetic age gap as an intermediate linking structural inequities to disparities in cognition and functioning with age. Historic and current manifestations of structural racism, discrimination, and segregation lead to inequities in education, income, and wealth between White and Black older Americans. Black Americans face higher epigenetic age gap partially as a result of lower SES which in turn contributes to increased risk of cognitive and functional impairment and decline.

The exact mechanisms by which low SES is associated with higher epigenetic age gap remain unclear, but likely relate to chronic stressors and reduced access to health-promoting resources (48). Other aspects of systemic racism, such as neighborhood-level disinvestment, environmental racism that results in higher exposure to pollutants for Black communities, and personal experiences of discrimination, likely contribute to this pathway in addition to inequities in individual SES (14,30,49,50). Further research is needed to elucidate these pathways and identify points of intervention.

Our results should be interpreted considering several strengths and limitations. The HRS DNAm sample is a large, diverse sample that is representative of Americans over 50. We incorporate sampling weights in all analyses to correct for potential sampling bias. We exclude participants who identify as Hispanic or any race other than Black or White due to smaller sample sizes. Mortality selection may result in underestimates of the epigenetic age gap associations because in this older sample Black participants who are biologically oldest and most impaired are more likely to have deceased. Although we include education, income, and wealth as measures of SES, we do not capture other aspects of SES such as childhood adversity, neighborhood deprivation or affluence, informal education, occupation and retirement, homeownership, and dependents or family support. We perform additional supplemental analyses with health variables thought to be important confounders of cognitive and functional outcomes, but the possibility of unmeasured or unadjusted confounding remains. Important future directions for research on disparities in aging are to include a greater diversity of racial, ethnic, language, and immigration groups as the older population becomes more socioculturally diverse and incoporating a life course approach to provide a greater understanding of how early-life conditions influence adult epigenetic, cognitive, and functional aging.

We use two independent outcome measures to strengthen our analyses and recognize that each outcome has weaknesses. For the longitudinal outcomes (change in TICS score and incidence of IADL limitations), we have only two years of follow-up data. This likely biases the results towards the null. TICS is a brief neuropsychological battery that has limited sensitivity and potential measurement bias and does not comprehensively probe all domains of cognition. Because of a skewed distribution of number of IADL limitations we use binary IADL prevalence and incidence outcomes which do not capture the degree of disability or decline. We are not able to assess causal relationships between epigenetic age gap and these outcomes due to the lack of longitudinal DNAm data. Raw DNAm data are also not yet available from HRS, which would allow us to incorporate next-generation epigenetic age measures such as DunedinPACE and investigate specific biologically-relevant DNAm sites and regions.

Eliminating the racial disparity in epigenetic age gap would likely greatly reduce disparities in many age-related health outcomes, including cognitive and IADL performance. Future longitudinal causal research is needed to test the effects of specific interventions on epigenetic age gap, and the effect of changing epigenetic age gap on outcomes. As inequitable levels of SES between older Black and White Americans are a driving contributor to disparities in both epigenetic age gap and many age-related outcomes, including cognition and functioning, interventions are needed to reduce or compensate for inequitable levels of education, wealth, and income.

## Data Availability

All data produced in the study are publicly available from the Health and Retirement Study. Code to reproduce the data set and analyses is available upon reasonable request to the authors.

https://hrsdata.isr.umich.edu/data-products/public-survey-data

## Acknowledgements

The authors thank Drs. Dawei Xie, Irma Elo, Lauren Massimo, and Eddie Lee for their input and feedback during the development and implementation of this project. All authors meet ICMJE criteria for authorship. IY made substantial contributions to conception, design, analysis, and interpretation and drafted the manuscript. SDS made substantial contributions to conception and interpretation. CB made substantial contributions to conception and interpretation. SXX made substantial contributions to design and interpretation of analyses. RTB made substantial contributions to conception, design, and acquisition of data. CTM made substantial contributions to conception, design, and interpretation. All authors reviewed the manuscript critically, approved its publication, and agree to be accountable for the work. The funders of this work had no role in creating of this manuscript.

