## Supplemental material for "Epigenetic age and socioeconomic status contribute to racial disparities in cognitive and functional aging between Black and White older Americans"

### Supplemental Methods

We perform supplemental regression models with health behavior and health status covariates that are potential confounders of the relationship between epigenetic age gap and cognitive functioning outcomes. We select variables that have been reported as risk factors for cognitive or IADL impairment and examine univariable and multivariable associations with epigenetic age gap and outcome measures. Health data are from the HRS survey in the 2014 wave. All measures are self-reported by the participant.

Health behavior variables include tobacco smoking, alcohol consumption, and frequency of physical activity (1). Tobacco smoking is coded as never smoked, smoked in the past, or currently smoke. Alcohol consumption is coded as none, moderate if participant reports drinking two or fewer drinks on days when they drink, or heavy if participant reports more than two drinks. Frequency of physical activity is reverse-coded so that a higher number indicates greater frequency and is reported on a five-point scale from never, 1-3 times per month, once per week, more than once per week, and every day. We take the higher value of either moderate or vigorous physical activity frequency.

Health status variables include self-rated health, number of depression symptoms, number of chronic conditions, and history of stroke. Self-rated health is reported on a five-point scale. We dichotomize the variable to fair/poor or good/very good/excellent. Depression symptoms are assessed using the Center for Epidemiologic Studies Depression (CESD) scale based on eight indicators (2). Number of depression symptoms is not significantly associated with epigenetic age gap in multivariable analyses, but we include it because it is an important risk factor for cognitive impairment and decline. Chronic conditions are high blood pressure, diabetes, cancer, lung disease, heart problems, and arthritis for a maximum of six. Participants were asked whether a doctor had ever diagnosed them with each condition. History of stroke is a binary variable indicating whether a doctor had ever told the participant they had a stroke. We considered including heart disease and diabetes as separate covariates, but neither were significantly associated with outcomes in multivariable analyses.

We additionally considered including measures of healthcare access, i.e. insurance status, whether the participant had delayed taking medications due to cost, and whether participant had visited a doctor in the past two years, as well as partnership status (married or partnered vs not) and region of birth. None of these were significantly associated with epigenetic age gap in univariable analyses. We previously demonstrated that adjusting for ancestry-informative principal component genetic markers did not change the distribution nor the racial disparity in epigenetic age gap and thus did not adjust for genetic ancestry in these analyses (3).

1.	Thierry AD, Sherman-Wilkins K, Armendariz M, Sullivan A, Farmer HR. Perceived Neighborhood Characteristics and Cognitive Functioning among Diverse Older Adults: An Intersectional Approach. *Int J Environ Res Public Health*. 2021;18(5):2661. doi:10.3390/ijerph18052661.

2.	Steffick D. *Documentation of Affective Functioning Measures in the Health and Retirement Study*. Ann Arbor, Michigan: Institute for Social Research, University of Michigan; 2000. doi:10.7826/ISR-UM.06.585031.001.05.0005.2000.

3.	Yannatos I, Stites S, Brown RT, McMillan CT. Contributions of neighborhood social environment and air pollution exposure to Black-White disparities in epigenetic aging. *PLOS ONE*. 2023;18(7):e0287112. doi:10.1371/journal.pone.0287112.

**Supplemental Tables**

**eTable 1: Basic and SES regression models with TICS score outcome**

| **Beta (95% CI)^1^** | **Basic^1^** | **SES^1^** | **Basic + GrimAge^1^** | **SES + GrimAge^1^** | **Basic + DPoAm^1^** | **SES + DPoAm^1^** |
| --- | --- | --- | --- | --- | --- | --- |
| **Age** | -0.16*** (-0.17,-0.14) | -0.13*** (-0.15,-0.12) | -0.15*** (-0.17,-0.14) | -0.13*** (-0.15,-0.12) | -0.15*** (-0.17,-0.14) | -0.13*** (-0.15,-0.12) |
| **Gender (Female)** | 0.89*** (0.62,1.2) | 1.0*** (0.79,1.3) | 0.35 (0.07,0.63) | 0.80*** (0.54,1.1) | 0.81*** (0.54,1.1) | 1.0*** (0.76,1.3) |
| **Race (Black)** | -3.1*** (-3.5,-2.6) | -1.9*** (-2.3,-1.5) | -2.7*** (-3.1,-2.3) | -1.8*** (-2.2,-1.4) | -2.9*** (-3.3,-2.5) | -1.8*** (-2.3,-1.4) |
| **Education** |  |  |  |  |  |  |
| College + |  | — |  | — |  | — |
| Some College |  | -1.2*** (-1.5,-0.87) |  | -1.1*** (-1.4,-0.78) |  | -1.2*** (-1.5,-0.84) |
| High School |  | -1.8*** (-2.2,-1.5) |  | -1.7*** (-2.0,-1.4) |  | -1.8*** (-2.1,-1.4) |
| < High School |  | -3.6*** (-4.1,-3.2) |  | -3.5*** (-4.0,-3.0) |  | -3.6*** (-4.1,-3.1) |
| **Wealth/Income Quartile** |  |  |  |  |  |  |
| 4 |  | — |  | — |  | — |
| 3 |  | -0.65*** (-0.99,-0.31) |  | -0.59** (-0.93,-0.25) |  | -0.65*** (-0.99,-0.31) |
| 2 |  | -1.0*** (-1.4,-0.65) |  | -0.89*** (-1.3,-0.53) |  | -0.98*** (-1.3,-0.62) |
| 1 |  | -2.0*** (-2.4,-1.6) |  | -1.8*** (-2.2,-1.4) |  | -1.9*** (-2.3,-1.5) |
| **Epigenetic age gap** |  |  | -0.77*** (-0.91,-0.64) | -0.33*** (-0.46,-0.19) | -0.47*** (-0.60,-0.33) | -0.15 (-0.28,-0.02) |
| **(Intercept)** | 26*** (25,27) | 27*** (26,28) | 27*** (26,28) | 27*** (26,28) | 26*** (25,27) | 27*** (26,28) |
| R² | 0.170 | 0.289 | 0.200 | 0.294 | 0.181 | 0.290 |
| AIC | 18,922 | 18,427 | 18,801 | 18,406 | 18,878 | 18,423 |
| No. Obs. | 3,282 | 3,282 | 3,282 | 3,282 | 3,282 | 3,282 |
| ^1^*p<0.05; **p<0.01; ***p<0.001 | | | | | | |

**eTable 2: Basic and SES regression models with TICS change outcome**

| **Beta (95% CI)^1^** | **Basic^1^** | **SES^1^** | **Basic + GrimAge^1^** | **SES + GrimAge^1^** | **Basic + DPoAm^1^** | **SES + DPoAm^1^** |
| --- | --- | --- | --- | --- | --- | --- |
| **Age** | -0.09*** (-0.10,-0.07) | -0.09*** (-0.10,-0.07) | -0.09*** (-0.10,-0.08) | -0.09*** (-0.10,-0.07) | -0.09*** (-0.10,-0.07) | -0.09*** (-0.10,-0.07) |
| **Gender (Female)** | 0.47*** (0.22,0.71) | 0.59*** (0.35,0.84) | 0.18 (-0.08,0.44) | 0.40* (0.14,0.66) | 0.43** (0.19,0.68) | 0.57*** (0.32,0.81) |
| **Race (Black)** | -1.8*** (-2.2,-1.4) | -1.4*** (-1.8,-1.0) | -1.6*** (-2.0,-1.3) | -1.4*** (-1.8,-0.98) | -1.7*** (-2.1,-1.3) | -1.4*** (-1.8,-0.99) |
| **TICS score** | -0.43*** (-0.47,-0.40) | -0.50*** (-0.53,-0.46) | -0.45*** (-0.49,-0.42) | -0.50*** (-0.54,-0.47) | -0.44*** (-0.47,-0.41) | -0.50*** (-0.53,-0.46) |
| **Education** |  |  |  |  |  |  |
| College + |  | — |  | — |  | — |
| Some College |  | -0.45* (-0.77,-0.13) |  | -0.38 (-0.70,-0.06) |  | -0.42* (-0.74,-0.10) |
| High School |  | -0.89*** (-1.2,-0.56) |  | -0.81*** (-1.1,-0.48) |  | -0.86*** (-1.2,-0.53) |
| < High School |  | -1.6*** (-2.1,-1.1) |  | -1.5*** (-2.0,-0.98) |  | -1.6*** (-2.1,-1.1) |
| **Wealth/Income Quartile** |  |  |  |  |  |  |
| 4 |  | — |  | — |  | — |
| 3 |  | -0.08 (-0.41,0.25) |  | -0.04 (-0.37,0.29) |  | -0.08 (-0.41,0.25) |
| 2 |  | -0.33 (-0.68,0.02) |  | -0.25 (-0.60,0.10) |  | -0.31 (-0.66,0.04) |
| 1 |  | -0.87*** (-1.3,-0.47) |  | -0.74** (-1.1,-0.33) |  | -0.82*** (-1.2,-0.41) |
| **Epigenetic age gap** |  |  | -0.42*** (-0.55,-0.29) | -0.26*** (-0.40,-0.13) | -0.26*** (-0.38,-0.13) | -0.14 (-0.27,-0.02) |
| **(Intercept)** | 13*** (12,15) | 15*** (14,16) | 14*** (13,15) | 15*** (14,16) | 13*** (12,15) | 15*** (14,16) |
| R² | 0.195 | 0.222 | 0.206 | 0.226 | 0.200 | 0.224 |
| AIC | 15,588 | 15,502 | 15,551 | 15,490 | 15,574 | 15,499 |
| No. Obs. | 2,849 | 2,849 | 2,849 | 2,849 | 2,849 | 2,849 |
| ^1^*p<0.05; **p<0.01; ***p<0.001 | | | | | | |

**eTable 3: Basic and SES regression models with IADL prevalence outcome**

| **Odds Ratio (95% CI)^1^** | **Basic^12^** | **SES^12^** | **Basic + GrimAge^12^** | **SES + GrimAge^12^** | **Basic + DPoAm^12^** | **SES + DPoAm^12^** |
| --- | --- | --- | --- | --- | --- | --- |
| **Age** | 1.05*** (1.04,1.06) | 1.04*** (1.03,1.06) | 1.05*** (1.04,1.07) | 1.05*** (1.03,1.06) | 1.05*** (1.04,1.06) | 1.05*** (1.03,1.06) |
| **Gender (Female)** | 1.18 (0.92,1.52) | 1.10 (0.85,1.43) | 1.66** (1.26,2.17) | 1.45* (1.10,1.92) | 1.24 (0.96,1.59) | 1.14 (0.88,1.48) |
| **Race (Black)** | 2.01*** (1.45,2.79) | 1.34 (0.92,1.93) | 1.69** (1.20,2.36) | 1.27 (0.88,1.84) | 1.83** (1.30,2.58) | 1.29 (0.89,1.88) |
| **Education** |  |  |  |  |  |  |
| College + |  | — |  | — |  | — |
| Some College |  | 1.66* (1.13,2.44) |  | 1.52 (1.03,2.24) |  | 1.61 (1.10,2.37) |
| High School |  | 1.93** (1.30,2.85) |  | 1.73* (1.17,2.56) |  | 1.85** (1.25,2.73) |
| < High School |  | 2.33** (1.46,3.72) |  | 1.97* (1.23,3.15) |  | 2.20** (1.38,3.52) |
| **Wealth/Income Quartile** |  |  |  |  |  |  |
| 4 |  | — |  | — |  | — |
| 3 |  | 0.73 (0.49,1.10) |  | 0.68 (0.45,1.01) |  | 0.73 (0.49,1.10) |
| 2 |  | 1.42 (0.98,2.07) |  | 1.27 (0.87,1.85) |  | 1.40 (0.96,2.04) |
| 1 |  | 1.94** (1.28,2.92) |  | 1.59 (1.05,2.42) |  | 1.84* (1.21,2.78) |
| **Epigenetic age gap** |  |  | 1.60*** (1.40,1.82) | 1.44*** (1.25,1.66) | 1.29*** (1.14,1.46) | 1.18* (1.04,1.34) |
| **(Intercept)** | 0.00*** (0.00,0.01) | 0.00*** (0.00,0.01) | 0.00*** (0.00,0.01) | 0.00*** (0.00,0.01) | 0.00*** (0.00,0.01) | 0.00*** (0.00,0.01) |
| Null deviance | 2,498 | 2,498 | 2,498 | 2,498 | 2,498 | 2,498 |
| AIC | 2,402 | 2,331 | 2,334 | 2,294 | 2,383 | 2,325 |
| No. Obs. | 3,282 | 3,282 | 3,282 | 3,282 | 3,282 | 3,282 |
| ^1^*p<0.05; **p<0.01; ***p<0.001 | | | | | | |
| ^2^OR = Odds Ratio | | | | | | |

**eTable 4: Basic and SES regression models with IADL incidence outcome**

| **Odds Ratio (95% CI)^1^** | **Basic^12^** | **SES^12^** | **Basic + GrimAge^12^** | **SES + GrimAge^12^** | **Basic + DPoAm^12^** | **SES + DPoAm^12^** |
| --- | --- | --- | --- | --- | --- | --- |
| **Age** | 1.06*** (1.04,1.08) | 1.05*** (1.03,1.07) | 1.06*** (1.04,1.08) | 1.06*** (1.03,1.08) | 1.06*** (1.03,1.08) | 1.05*** (1.03,1.07) |
| **Gender (Female)** | 1.12 (0.78,1.61) | 1.07 (0.74,1.54) | 1.69* (1.14,2.50) | 1.55 (1.03,2.31) | 1.21 (0.84,1.74) | 1.16 (0.80,1.66) |
| **Race (Black)** | 2.02* (1.27,3.23) | 1.28 (0.77,2.13) | 1.65 (1.00,2.71) | 1.20 (0.72,2.02) | 1.64 (1.02,2.64) | 1.14 (0.69,1.89) |
| **Education** |  |  |  |  |  |  |
| College + |  | — |  | — |  | — |
| Some College |  | 1.15 (0.66,2.00) |  | 1.00 (0.57,1.76) |  | 1.05 (0.61,1.83) |
| High School |  | 1.18 (0.68,2.04) |  | 1.01 (0.58,1.75) |  | 1.06 (0.62,1.82) |
| < High School |  | 1.71 (0.84,3.49) |  | 1.40 (0.67,2.93) |  | 1.50 (0.73,3.06) |
| **Wealth/Income Quartile** |  |  |  |  |  |  |
| 4 |  | — |  | — |  | — |
| 3 |  | 1.09 (0.62,1.91) |  | 0.99 (0.56,1.74) |  | 1.09 (0.62,1.90) |
| 2 |  | 1.86 (1.05,3.29) |  | 1.64 (0.92,2.94) |  | 1.79 (1.01,3.17) |
| 1 |  | 2.72* (1.41,5.25) |  | 2.15 (1.08,4.27) |  | 2.42* (1.22,4.79) |
| **Epigenetic age gap** |  |  | 1.72*** (1.44,2.06) | 1.59*** (1.31,1.93) | 1.57*** (1.30,1.90) | 1.48*** (1.22,1.79) |
| **(Intercept)** | 0.00*** (0.00,0.01) | 0.00*** (0.00,0.01) | 0.00*** (0.00,0.00) | 0.00*** (0.00,0.00) | 0.00*** (0.00,0.01) | 0.00*** (0.00,0.00) |
| Null deviance | 1,284 | 1,284 | 1,284 | 1,284 | 1,284 | 1,284 |
| AIC | 1,241 | 1,224 | 1,201 | 1,199 | 1,211 | 1,204 |
| No. Obs. | 2,538 | 2,538 | 2,538 | 2,538 | 2,538 | 2,538 |
| ^1^*p<0.05; **p<0.01; ***p<0.001 | | | | | | |
| ^2^OR = Odds Ratio | | | | | | |

**eTable 5: Health characteristics by race**

| **Characteristic** | **Overall, N = 3,282^1^** | **White, N = 2,636^1^** | **Black, N = 646^1^** | **p-value^2^** | **Effect Size^3^** |
| --- | --- | --- | --- | --- | --- |
| **Tobacco smoking** |  |  |  | <0.001 | 0.102 |
| Never | 1,410 (43%) | 1,144 (44%) | 266 (41%) |  |  |
| Past | 1,443 (44%) | 1,190 (45%) | 253 (39%) |  |  |
| Current | 410 (13%) | 286 (11%) | 124 (19%) |  |  |
| Unknown | 19 | 16 | 3 |  |  |
| **Alcohol consumption** |  |  |  | <0.001 | 0.103 |
| None | 1,390 (42%) | 1,053 (40%) | 337 (52%) |  |  |
| Moderate | 1,580 (48%) | 1,333 (51%) | 247 (38%) |  |  |
| Heavy | 301 (9.2%) | 239 (9.1%) | 62 (9.6%) |  |  |
| Unknown | 11 | 11 | 0 |  |  |
| **Physical activity frequency** | 2.17 (1.27) | 2.23 (1.27) | 1.94 (1.28) | <0.001 | 0.222 |
| **Self-rated health** |  |  |  | <0.001 | 0.118 |
| Good/Great | 2,554 (78%) | 2,117 (80%) | 437 (68%) |  |  |
| Fair/Poor | 726 (22%) | 519 (20%) | 207 (32%) |  |  |
| Unknown | 2 | 0 | 2 |  |  |
| **Depression symptoms** | 1.31 (1.92) | 1.19 (1.85) | 1.79 (2.09) | <0.001 | 0.305 |
| Unknown | 10 | 8 | 2 |  |  |
| **# Chronic conditions** | 1.97 (1.28) | 1.94 (1.28) | 2.13 (1.27) | <0.001 | 0.151 |
| **Stroke** |  |  |  | 0.021 | 0.039 |
| 0 | 3,010 (92%) | 2,432 (92%) | 578 (89%) |  |  |
| 1 | 272 (8.3%) | 204 (7.7%) | 68 (11%) |  |  |
| ^1^n (%); Mean (SD) | | | | | |
| ^2^Pearson's Chi-squared test; Wilcoxon rank sum test | | | | | |
| ^3^Cohen's D; Cramer's V | | | | | |

**eTable 6: Health behavior and status models for TICS outcomes**

|  | **GrimAge** | | | | **DPoAm** | | | |
| --- | --- | --- | --- | --- | --- | --- | --- | --- |
|  | **TICS Score** | | **TICS Change** | | **TICS Score** | | **TICS Change** | |
| **Beta (95% CI)^1^** | **+ Behaviors^1^** | **+ Health Status^1^** | **+ Behaviors^1^** | **+ Health Status^1^** | **+ Behaviors^1^** | **+ Health Status^1^** | **+ Behaviors^1^** | **+ Health Status^1^** |
| **Epigenetic age gap** | -0.29*** (-0.45,-0.14) | -0.25** (-0.41,-0.09) | -0.25** (-0.41,-0.10) | -0.22* (-0.38,-0.06) | -0.09 (-0.22,0.05) | -0.07 (-0.21,0.06) | -0.11 (-0.25,0.02) | -0.09 (-0.23,0.05) |
| **Age** | -0.13*** (-0.14,-0.12) | -0.13*** (-0.15,-0.12) | -0.09*** (-0.10,-0.07) | -0.09*** (-0.11,-0.08) | -0.13*** (-0.14,-0.12) | -0.13*** (-0.15,-0.12) | -0.09*** (-0.10,-0.07) | -0.09*** (-0.11,-0.08) |
| **Gender (Female)** | 0.85*** (0.58,1.1) | 0.88*** (0.61,1.2) | 0.40* (0.13,0.67) | 0.43** (0.16,0.71) | 1.0*** (0.78,1.3) | 1.0*** (0.78,1.3) | 0.55*** (0.30,0.81) | 0.56*** (0.31,0.82) |
| **Race (Black)** | -1.8*** (-2.2,-1.4) | -1.8*** (-2.2,-1.4) | -1.4*** (-1.8,-0.98) | -1.4*** (-1.8,-0.97) | -1.9*** (-2.3,-1.5) | -1.8*** (-2.2,-1.4) | -1.4*** (-1.8,-1.0) | -1.4*** (-1.8,-0.98) |
| **Education** |  |  |  |  |  |  |  |  |
| College + | — | — | — | — | — | — | — | — |
| Some College | -1.0*** (-1.3,-0.68) | -1.0*** (-1.4,-0.71) | -0.34 (-0.66,-0.01) | -0.34 (-0.66,-0.02) | -1.1*** (-1.4,-0.72) | -1.1*** (-1.4,-0.73) | -0.36 (-0.68,-0.03) | -0.36 (-0.68,-0.04) |
| High School | -1.6*** (-1.9,-1.3) | -1.6*** (-1.9,-1.2) | -0.78*** (-1.1,-0.45) | -0.81*** (-1.1,-0.47) | -1.7*** (-2.0,-1.3) | -1.6*** (-1.9,-1.3) | -0.81*** (-1.2,-0.48) | -0.84*** (-1.2,-0.50) |
| < High School | -3.3*** (-3.8,-2.8) | -3.2*** (-3.7,-2.8) | -1.5*** (-2.0,-0.95) | -1.4*** (-2.0,-0.93) | -3.4*** (-3.9,-2.9) | -3.3*** (-3.8,-2.8) | -1.5*** (-2.0,-0.99) | -1.5*** (-2.0,-0.97) |
| **Wealth/Income Quartile** |  |  |  |  |  |  |  |  |
| 4 | — | — | — | — | — | — | — | — |
| 3 | -0.54** (-0.89,-0.20) | -0.55** (-0.89,-0.22) | -0.01 (-0.34,0.32) | 0.01 (-0.32,0.34) | -0.58** (-0.92,-0.24) | -0.58** (-0.92,-0.24) | -0.04 (-0.37,0.29) | -0.01 (-0.34,0.32) |
| 2 | -0.75*** (-1.1,-0.39) | -0.69*** (-1.1,-0.32) | -0.19 (-0.55,0.17) | -0.12 (-0.48,0.24) | -0.80*** (-1.2,-0.43) | -0.72*** (-1.1,-0.35) | -0.22 (-0.58,0.14) | -0.14 (-0.50,0.22) |
| 1 | -1.6*** (-2.0,-1.2) | -1.4*** (-1.8,-0.98) | -0.68** (-1.1,-0.26) | -0.48 (-0.91,-0.06) | -1.7*** (-2.1,-1.3) | -1.4*** (-1.9,-1.0) | -0.73** (-1.1,-0.31) | -0.52 (-0.94,-0.10) |
| **Tobacco smoking** |  |  |  |  |  |  |  |  |
| Never | — | — | — | — | — | — | — | — |
| Past | -0.06 (-0.33,0.22) | -0.03 (-0.31,0.24) | 0.04 (-0.23,0.31) | 0.06 (-0.21,0.32) | -0.15 (-0.42,0.12) | -0.11 (-0.38,0.16) | -0.02 (-0.29,0.24) | 0.00 (-0.27,0.26) |
| Current | -0.04 (-0.51,0.43) | 0.04 (-0.43,0.51) | 0.03 (-0.44,0.50) | 0.07 (-0.40,0.54) | -0.38 (-0.82,0.07) | -0.23 (-0.67,0.21) | -0.20 (-0.65,0.24) | -0.14 (-0.59,0.30) |
| **Alcohol consumption** |  |  |  |  |  |  |  |  |
| None | — | — | — | — | — | — | — | — |
| Moderate | 0.53*** (0.25,0.81) | 0.48** (0.20,0.75) | 0.23 (-0.04,0.51) | 0.19 (-0.09,0.47) | 0.56*** (0.28,0.84) | 0.49** (0.22,0.77) | 0.25 (-0.02,0.53) | 0.21 (-0.07,0.48) |
| Heavy | 0.49 (0.05,0.94) | 0.42 (-0.02,0.86) | 0.00 (-0.43,0.43) | -0.03 (-0.46,0.40) | 0.48 (0.04,0.93) | 0.40 (-0.04,0.84) | -0.01 (-0.44,0.42) | -0.04 (-0.48,0.39) |
| **Physical activity frequency** | 0.10 (0.00,0.21) | 0.04 (-0.07,0.14) | 0.02 (-0.09,0.12) | -0.05 (-0.15,0.06) | 0.12 (0.01,0.22) | 0.05 (-0.06,0.15) | 0.03 (-0.07,0.13) | -0.04 (-0.14,0.07) |
| **Self-rated health** |  |  |  |  |  |  |  |  |
| Good/Great |  | — |  | — |  | — |  | — |
| Fair/Poor |  | -0.62** (-0.98,-0.27) |  | -0.42 (-0.78,-0.06) |  | -0.66** (-1.0,-0.30) |  | -0.45 (-0.81,-0.09) |
| **Depression symptoms** |  | -0.12** (-0.19,-0.05) |  | -0.11** (-0.18,-0.04) |  | -0.12** (-0.19,-0.05) |  | -0.11* (-0.18,-0.04) |
| **# Chronic conditions** |  | 0.06 (-0.05,0.17) |  | -0.02 (-0.13,0.09) |  | 0.05 (-0.06,0.16) |  | -0.03 (-0.14,0.08) |
| **Stroke** |  |  |  |  |  |  |  |  |
| 0 |  | — |  | — |  | — |  | — |
| 1 |  | -0.68* (-1.2,-0.18) |  | -0.43 (-0.95,0.10) |  | -0.71* (-1.2,-0.21) |  | -0.45 (-0.97,0.08) |
| **TICS score** |  |  | -0.51*** (-0.54,-0.47) | -0.52*** (-0.55,-0.48) |  |  | -0.50*** (-0.54,-0.47) | -0.51*** (-0.55,-0.48) |
| **(Intercept)** | 26*** (25,27) | 26*** (25,27) | 15*** (13,16) | 16*** (14,17) | 26*** (25,27) | 26*** (25,27) | 15*** (13,16) | 16*** (14,17) |
| R² | 0.298 | 0.309 | 0.230 | 0.237 | 0.295 | 0.307 | 0.228 | 0.236 |
| AIC | 18,222 | 18,094 | 15,355 | 15,313 | 18,234 | 18,102 | 15,363 | 15,318 |
| No. Obs. | 3,252 | 3,240 | 2,825 | 2,821 | 3,252 | 3,240 | 2,825 | 2,821 |
| ^1^*p<0.05; **p<0.01; ***p<0.001 | | | | | | | | |

**eTable 7: Health behavior and status models for IADL outcomes**

|  | **GrimAge** | | | | **DPoAm** | | | |
| --- | --- | --- | --- | --- | --- | --- | --- | --- |
|  | **Prevalence** | | **Incidence** | | **Prevalence** | | **Incidence** | |
| **Odds Ratio (95% CI)^1^** | **+ Behaviors^12^** | **+ Health Status^12^** | **+ Behaviors^12^** | **+ Health Status^12^** | **+ Behaviors^12^** | **+ Health Status^12^** | **+ Behaviors^12^** | **+ Health Status^12^** |
| **Epigenetic age gap** | 1.36** (1.14,1.61) | 1.20 (0.99,1.46) | 1.56*** (1.24,1.96) | 1.48** (1.17,1.88) | 1.05 (0.92,1.21) | 1.00 (0.86,1.16) | 1.41** (1.14,1.74) | 1.36* (1.11,1.66) |
| **Age** | 1.04*** (1.02,1.05) | 1.04*** (1.03,1.06) | 1.06*** (1.03,1.08) | 1.06*** (1.03,1.09) | 1.04*** (1.03,1.06) | 1.04*** (1.03,1.06) | 1.06*** (1.03,1.08) | 1.06*** (1.03,1.08) |
| **Gender (Female)** | 1.20 (0.89,1.63) | 1.15 (0.85,1.57) | 1.51 (0.98,2.32) | 1.49 (0.96,2.31) | 0.99 (0.74,1.31) | 1.03 (0.76,1.39) | 1.16 (0.79,1.69) | 1.18 (0.79,1.74) |
| **Race (Black)** | 1.29 (0.89,1.88) | 1.24 (0.84,1.84) | 1.18 (0.69,2.02) | 1.14 (0.67,1.95) | 1.34 (0.91,1.96) | 1.28 (0.86,1.91) | 1.14 (0.68,1.90) | 1.09 (0.64,1.83) |
| **Education** |  |  |  |  |  |  |  |  |
| College + | — | — | — | — | — | — | — | — |
| Some College | 1.37 (0.92,2.04) | 1.29 (0.85,1.96) | 0.91 (0.51,1.63) | 0.88 (0.49,1.58) | 1.42 (0.96,2.11) | 1.31 (0.87,1.99) | 0.95 (0.54,1.67) | 0.91 (0.51,1.61) |
| High School | 1.51 (1.01,2.26) | 1.54 (1.02,2.31) | 0.92 (0.53,1.62) | 0.94 (0.54,1.64) | 1.57 (1.05,2.35) | 1.57 (1.05,2.36) | 0.95 (0.55,1.66) | 0.95 (0.55,1.66) |
| < High School | 1.64 (1.01,2.68) | 1.50 (0.90,2.49) | 1.20 (0.57,2.52) | 1.11 (0.53,2.34) | 1.73 (1.06,2.83) | 1.55 (0.93,2.58) | 1.23 (0.59,2.56) | 1.12 (0.53,2.35) |
| **Wealth/Income Quartile** |  |  |  |  |  |  |  |  |
| 4 | — | — | — | — | — | — | — | — |
| 3 | 0.62 (0.41,0.93) | 0.54* (0.35,0.82) | 0.88 (0.50,1.56) | 0.79 (0.44,1.43) | 0.64 (0.42,0.96) | 0.54* (0.35,0.83) | 0.96 (0.54,1.67) | 0.84 (0.47,1.52) |
| 2 | 1.02 (0.68,1.52) | 0.81 (0.54,1.22) | 1.41 (0.76,2.59) | 1.24 (0.66,2.31) | 1.06 (0.71,1.57) | 0.83 (0.55,1.24) | 1.47 (0.80,2.71) | 1.29 (0.69,2.42) |
| 1 | 1.25 (0.80,1.95) | 0.77 (0.48,1.23) | 1.79 (0.88,3.65) | 1.28 (0.62,2.65) | 1.34 (0.86,2.09) | 0.79 (0.49,1.28) | 1.93 (0.94,3.96) | 1.39 (0.67,2.88) |
| **Tobacco smoking** |  |  |  |  |  |  |  |  |
| Never | — | — | — | — | — | — | — | — |
| Past | 1.04 (0.77,1.42) | 0.96 (0.70,1.32) | 0.91 (0.61,1.35) | 0.88 (0.59,1.32) | 1.18 (0.88,1.58) | 1.03 (0.76,1.40) | 1.00 (0.68,1.47) | 0.95 (0.64,1.41) |
| Current | 1.05 (0.64,1.72) | 1.01 (0.59,1.71) | 0.96 (0.48,1.93) | 0.90 (0.45,1.79) | 1.56 (1.01,2.42) | 1.31 (0.81,2.11) | 1.24 (0.65,2.35) | 1.09 (0.57,2.09) |
| **Alcohol consumption** |  |  |  |  |  |  |  |  |
| None | — | — | — | — | — | — | — | — |
| Moderate | 0.64** (0.48,0.85) | 0.72 (0.53,0.97) | 0.60 (0.40,0.91) | 0.65 (0.43,0.98) | 0.62** (0.46,0.82) | 0.71 (0.53,0.96) | 0.59* (0.39,0.88) | 0.64 (0.43,0.97) |
| Heavy | 0.47* (0.27,0.80) | 0.54 (0.30,0.98) | 1.17 (0.62,2.23) | 1.25 (0.65,2.38) | 0.47* (0.28,0.81) | 0.55 (0.30,0.99) | 1.20 (0.63,2.27) | 1.29 (0.68,2.45) |
| **Physical activity frequency** | 0.70*** (0.63,0.78) | 0.81*** (0.73,0.91) | 0.88 (0.75,1.02) | 0.98 (0.84,1.13) | 0.69*** (0.62,0.76) | 0.81*** (0.72,0.90) | 0.87 (0.74,1.01) | 0.96 (0.83,1.12) |
| **Self-rated health** |  |  |  |  |  |  |  |  |
| Good/Great |  | — |  | — |  | — |  | — |
| Fair/Poor |  | 2.12*** (1.53,2.94) |  | 2.12** (1.33,3.37) |  | 2.16*** (1.56,3.01) |  | 2.18** (1.37,3.46) |
| **Depression symptoms** |  | 1.26*** (1.18,1.34) |  | 1.17** (1.07,1.28) |  | 1.25*** (1.18,1.34) |  | 1.16** (1.06,1.27) |
| **# Chronic conditions** |  | 1.26*** (1.13,1.40) |  | 1.21 (1.03,1.43) |  | 1.28*** (1.16,1.42) |  | 1.22 (1.04,1.44) |
| **Stroke** |  |  |  |  |  |  |  |  |
| 0 |  | — |  | — |  | — |  | — |
| 1 |  | 1.61* (1.12,2.32) |  | 1.34 (0.65,2.75) |  | 1.65* (1.15,2.37) |  | 1.41 (0.70,2.88) |
| **(Intercept)** | 0.02*** (0.00,0.05) | 0.00*** (0.00,0.01) | 0.00*** (0.00,0.01) | 0.00*** (0.00,0.00) | 0.02*** (0.00,0.05) | 0.00*** (0.00,0.01) | 0.00*** (0.00,0.01) | 0.00*** (0.00,0.00) |
| Null deviance | 2,478 | 2,465 | 1,274 | 1,271 | 2,478 | 2,465 | 1,274 | 1,271 |
| AIC | 2,193 | 1,986 | 1,187 | 1,140 | 2,211 | 1,991 | 1,190 | 1,142 |
| No. Obs. | 3,252 | 3,240 | 2,518 | 2,513 | 3,252 | 3,240 | 2,518 | 2,513 |
| ^1^*p<0.05; **p<0.01; ***p<0.001 | | | | | | | | |
| ^2^OR = Odds Ratio | | | | | | | | |

**eTable 8: Full mediation results.**

|  | **Mediator:** | **GrimAge** | | **DPoAm** | |
| --- | --- | --- | --- | --- | --- |
|  | **Model:** | **Basic** | **SES** | **Basic** | **SES** |
| **Outcome** | **Effect** |  |  |  |  |
| TICS Score | Controlled Direct | -2.73*** (-3.17, -2.27) | -2.11*** (-2.56, -1.63) | -2.87*** (-3.29, -2.49) | -2.16*** (-2.63, -1.67) |
|  | Pure Natural Direct | -2.73*** (-3.17, -2.27) | -2.11*** (-2.56, -1.63) | -2.87*** (-3.29, -2.49) | -2.16*** (-2.63, -1.67) |
|  | Total Natural Direct | -2.73*** (-3.17, -2.27) | -2.11*** (-2.56, -1.63) | -2.87*** (-3.29, -2.49) | -2.16*** (-2.63, -1.67) |
|  | Pure Natural Indirect | -0.32*** (-0.43, -0.21) | -0.06*** (-0.13, -0.03) | -0.19*** (-0.28, -0.11) | -0.44 (-1.03, 0.73) |
|  | Total Natural Indirect | -0.32*** (-0.43, -0.21) | -0.06*** (-0.13, -0.03) | -0.19*** (-0.28, -0.11) | -0.44 (-1.03, 0.73) |
|  | Total | -3.06*** (-3.49, -2.58) | -2.17*** (-2.63, -1.68) | -3.06*** (-3.47, -2.70) | -2.60*** (-3.13, -1.31) |
|  | Mediated | 0.11*** (0.07, 0.14) | 0.03*** (0.01, 0.06) | 0.06*** (0.04, 0.09) | 0.17 (-0.54, 0.36) |
| TICS Change | Controlled Direct | -1.32*** (-1.70, -0.97) | -1.06*** (-1.57, -0.61) | -1.38*** (-1.82, -1.04) | -1.08*** (-1.58, -0.65) |
|  | Pure Natural Direct | -1.32*** (-1.70, -0.97) | -1.06*** (-1.57, -0.61) | -1.38*** (-1.82, -1.04) | -1.08*** (-1.58, -0.65) |
|  | Total Natural Direct | -1.32*** (-1.70, -0.97) | -1.06*** (-1.57, -0.61) | -1.38*** (-1.82, -1.04) | -1.08*** (-1.58, -0.65) |
|  | Pure Natural Indirect | -0.16*** (-0.26, -0.08) | 0.20 (-1.05, 1.10) | -0.10*** (-0.16, -0.02) | -0.29 (-0.78, 0.67) |
|  | Total Natural Indirect | -0.16*** (-0.26, -0.08) | 0.20 (-1.05, 1.10) | -0.10*** (-0.16, -0.02) | -0.29 (-0.78, 0.67) |
|  | Total | -1.48*** (-1.83, -1.10) | -0.86 (-2.12, 0.35) | -1.48*** (-1.89, -1.11) | -1.37* (-1.94, -0.32) |
|  | Mediated | 0.11*** (0.05, 0.18) | -0.23 (-2.52, 3.21) | 0.07*** (0.01, 0.11) | 0.21 (-1.31, 0.49) |
| IADL Prevalence | Controlled Direct | 1.65* (1.16, 2.21) | 1.20 (0.84, 1.73) | 1.79*** (1.32, 2.36) | 1.27 (0.82, 1.85) |
|  | Pure Natural Direct | 1.63* (1.16, 2.14) | 1.20 (0.85, 1.72) | 1.79*** (1.32, 2.33) | 1.26 (0.82, 1.85) |
|  | Total Natural Direct | 1.63* (1.16, 2.13) | 1.19 (0.85, 1.72) | 1.78*** (1.32, 2.32) | 1.27 (0.82, 1.85) |
|  | Pure Natural Indirect | 1.20*** (1.14, 1.28) | 1.08*** (1.04, 1.14) | 1.10*** (1.05, 1.17) | 1.04* (1.01, 1.08) |
|  | Total Natural Indirect | 1.20*** (1.13, 1.27) | 1.07*** (1.03, 1.14) | 1.10*** (1.05, 1.16) | 1.04* (1.01, 1.08) |
|  | Total | 1.96*** (1.35, 2.60) | 1.29 (0.90, 1.84) | 1.97*** (1.45, 2.63) | 1.31 (0.87, 1.87) |
|  | Mediated | 0.34*** (0.23, 0.58) | 0.31 (-0.90, 2.09) | 0.19*** (0.09, 0.32) | 0.16 (-0.83, 1.18) |
| IADL Incidence | Controlled Direct | 1.62 (0.98, 2.56) | 1.20 (0.66, 1.96) | 1.62 (0.91, 2.39) | 1.17 (0.67, 2.00) |
|  | Pure Natural Direct | 1.61 (0.98, 2.53) | 1.19 (0.67, 1.93) | 1.61 (0.91, 2.35) | 1.17 (0.67, 1.98) |
|  | Total Natural Direct | 1.61 (0.98, 2.52) | 1.19 (0.67, 1.93) | 1.60 (0.92, 2.35) | 1.17 (0.68, 1.98) |
|  | Pure Natural Indirect | 1.25*** (1.15, 1.38) | 1.12*** (1.05, 1.23) | 1.24* (1.11, 1.39) | 1.15*** (1.06, 1.27) |
|  | Total Natural Indirect | 1.24*** (1.15, 1.37) | 1.12*** (1.05, 1.23) | 1.24* (1.11, 1.38) | 1.15*** (1.06, 1.27) |
|  | Total | 2.01*** (1.21, 3.03) | 1.34 (0.80, 2.09) | 1.99* (1.17, 2.99) | 1.34 (0.73, 2.25) |
|  | Mediated | 0.39*** (0.23, 1.11) | 0.43 (-1.51, 10.44) | 0.39* (0.18, 0.85) | 0.50 (-2.78, 2.65) |
| *p<0.05; **p<0.01; ***p<0.001; (95% Confidence Intervals) | | | | | |
